# Association between genes regulating neural pathways for quantitative traits of speech and language disorders

**DOI:** 10.1101/2021.02.09.21251441

**Authors:** Penelope Benchek, Robert P Igo, Heather Voss-Hoynes, Yvonne Wren, Gabrielle Miller, Barbara Truitt, Wen Zhang, Michael Osterman, Lisa Freebairn, Jessica Tag, H. Gerry Taylor, E. Ricky Chan, Panos Roussos, Barbara Lewis, Catherine M. Stein, Sudha K. Iyengar

## Abstract

Speech sound disorders (SSD) manifest as difficulties in phonological memory and awareness, oral motor function, language, vocabulary, reading and spelling. Families enriched for SSD are rare, and typically display a cluster of deficits. We conducted a genome-wide association study (GWAS) in 435 children from 148 families in the Cleveland Family Speech and Reading study (CFSRS), examining 16 variables representing 6 domains. Replication was conducted using the Avon Longitudinal Study of Parents and Children (ALSPAC). We identified 18 significant loci (combined p<10^−8^) that we pursued bioinformatically. We prioritized 5 novel gene regions with likely functional repercussions on neural pathways, some which colocalized with differentially methylated regions in our sample. Polygenic risk scores for receptive language, expressive vocabulary, phonological awareness, phonological memory, spelling, and reading decoding associated with increasing clinical severity. In summary, neural genetic influence on SSD is primarily multigenic and acts on genomic regulatory elements, similar to other neurodevelopmental disorders.

## INTRODUCTION

Communication disorders are highly prevalent in the United States with approximately one in twelve children ages 3-17 years demonstrating a disorder ^1^. The most common difficulties are a speech problem (5%) or language problem (3.3%). Speech Sound disorders (SSD) include both errors of articulation or phonetic structure (errors due to poor motor abilities associated with the production of speech sounds) and phonological errors (errors in applying linguistic rules to combine sounds to form words). SSD have a prevalence of approximately 16% in children 3 years. of age^2^, with an estimated 3.8% of children persisting with speech delay at 6 years of age^3^. More than half of these children encounter later academic difficulties in language, reading, and spelling^7-11^. Because of the relative rarity of persistent speech problems and their correlation with other communication domains, endophenotypes are key to the study of genetic underpinnings.

Vocabulary is core to speech acquisition^4^. Children with difficulties in speech sound development often have difficulties with oral language and later reading and spelling disability^2,5-8^. Thus, speech, language, reading, and spelling measures are highly correlated and often have common genetic associations^9,10^. Moreover, speech and other communication phenotypes follow a developmental trajectory, where some speech and language disorders resolve with age, whereas others persist; genetic influences on the less easily resolved manifestations are generally stronger^11,12^. Because of the common genetic underpinnings and pathologic associations between speech and other communication phenotypes, it is conceivable that genetic replication interweaves with different communication measures. Of 7 known GWASs, none overlap in their top results (at p<5×10^−5^, see Table 3 ^13^), because they only focused on a limited number of phenotypes, or these measures were assessed at different ages (either pre-school or early school-age) ^13-20^, they only present results from one or a few measures and/or a binary trait; thus, the complexity of shared genetic influences is poorly understood. Most have not focused on children with SSD, particularly measures of articulation. Our sample represents a unique set of deeply phenotyped individuals with information on 6 domains that form the core of speech and language.

SSD are likely due to deficits in both motor ability and broader neural dysfunction. While motor deficits contribute to problems in speech production, abnormalities in other neural systems likely influence formation of phonological representation, which is common to SSD as well as reading and language impairment. We hypothesize that genetic regulation of these neural pathways is associated with variation common to speech, language, reading, and spelling ability. We conducted a GWAS in the Cleveland Family Speech and Reading Study (CFSRS), a cohort ascertained through a proband with SSD. We also conducted a methylome-wide study (i.e. MWAS) to determine the functional implications of these genetic associations, and replicated findings in a population-based cohort. We utilized a family-based cohort as our discovery sample because we hypothesized it would be enriched for disease-associated variants^21,22^. In these analyses, we identified new candidate genes for correlated communication endophenotypes, and bioinformatic annotation of these loci revealed that regulation of neural pathways is associated with variation in these measures.

## SUBJECTS AND METHODS

### Subject ascertainment – Cleveland Family Speech and Reading Study

From the Cleveland Family Speech and Reading Study (CFSRS)^23-28^, we examined 435 individuals from 148 families who had both DNA and endophenotype data available (Table 1). As previously described, families were ascertained through a proband with SSD identified from caseloads of speech-language pathologists in the Greater Cleveland area and referred to the study; detailed inclusion criteria are provided in the Supplemental Methods. Diagnosis of CAS was confirmed by an experienced licensed speech-language pathologist upon enrollment into the study. Socioeconomic status was determined at the initial assessment based on parent education levels and occupations using the Hollingshead Four Factor Index of Social Class^29^. This study was approved by the Institutional Review Board of Case Medical Center and University Hospitals and all parents provided informed consent and children older than 5 years provided assent.

**Table 1.** Characteristic table for CFSRS GWAS sample

|  |  |
| --- | --- |
| N* | 435 |
| Number of Families | 148 |
| Age range | [2.5, 64] |
| Female N (%) | 194 (45%) |
| Speech Disorder Subgroup N (%) |  |
| CAS | 47 (11%) |
| SSD + LI (no CAS) | 70 (16%) |
| SSD only | 119 (27%) |
| Lang only | 17 (4%) |
| No CAS/SSD/Lang | 177 (41%) |
| Missing | 5 (1%) |
| Hollingshead SES |  |
| 1 (lowest) | 3 (1%) |
| 2 | 30 (7%) |
| 3 | 67 (15%) |
| 4 | 167 (38%) |
| 5 (highest) | 147 (34%) |
| Missing | 21 (5%) |

\*Sample considered is the union of all samples across the 16 tests. Specific test sample sizes and age ranges are shown in supplemental Table 1.

### Communication Measures in CFSRS

We examined diadochokinetic rates using the *Robbins and Klee Oral Speech Motor Control Protocol* ^30^ or *Fletcher Time-by-Count Test of Diadochokinetic Syllable Rate*^*31*^. The merged variable is referred to as DDK. Expressive vocabulary was assessed with the *Expressive One Word Picture Vocabulary Test-Revised* (*EOWPVT*^*32*^) and receptive vocabulary with the *Peabody Picture Vocabulary Test-Third Edition (PPVT*^*33*^), and phonological memory with the *Nonsense Word Repetition (NSW* ^*34*^*), Multisyllabic Word Repetition (MSW* ^*34*^*), and Rapid Color Naming* ^*35*^ task. In addition to examining the total number of words correct for the MSW and NSW, we also examined the percent phonemes correct for both of these tasks (NSW-PPC and MSW-PPC, respectively). Phonological awareness was assessed using the *Elision* subtest of the *Comprehensive Test of Phonological Processing* – 2^nd^ Edition^36^. Reading was assessed using the *Woodcock Reading Mastery Test-Revised, Word Attack subtest (WRMT-AT*) and *Word Identification Subtest (WRMT-ID), the Reading Comprehension subtest (WIAT-RC)* and *Listening Comprehension subtest (WIAT-LC)* of the *Wechsler Individual Achievement Test* ^*37*^ Spelling was assessed on the *Test of Written Spelling-3 (TWS)* using the total score^38^.

Expressive and receptive language were assessed using the *Test of Language Development (TOLD*^*39*^) and *Clinical Evaluation of Language Fundamentals-Revised (CELF*^*40*^). referred to as the CELF-E (expressive) and CELF-R (receptive), respectively. Additional details about these measures are provided in the Supplemental Methods. For each of our tests we selected the first available assessment for each individual (Supplemental Table 1).

### GWAS analysis

Genotyping methods and quality control (QC) are described in the Supplemental Methods. Principal components (PC) obtained from principal component analysis (PCA) and the genetic relationship matrix (GRM) were generated using genotyped markers that met QC criteria. We used PC-AiR and PC-Relate from the Bioconductor package GENESIS^41^ to generate our PCs and GRM, respectively. PC-AiR accounts for sample relatedness to provide ancestry inference that is not confounded by family structure, while PC-Relate uses the ancestry representative PCs from PC-AiR to provide relatedness estimates due only to recent family (pedigree) structure.

To examine cross-trait correlation, we used GCTA^42^ to run a bivariate REML analysis for each pair of tests and tested for genetic correlations equal to 0. GCTA’s bivariate REML analysis estimates the genetic variance of each test and the genetic covariance between the two tests that can be captured by all SNPs^43^. Here we included all SNPs with MAF ≥ 0.01. The genetic variance/covariance calculated was adjusted for sex and the first two PCs.

We used RVTests, version 2.0^44^ to run our GWAS. We specifically relied on RVTest’s Grammar-gamma test^45^, which performs a linear mixed model association test while allowing for genotype dosages and accounting for family structure using the Genetic Relationship Matrix (GRM). Because each of our tests were age-normed we included only sex and the first two PCs as covariates in our regression models.

In addition, we generated endophenotype-based polygenic risk scores (PRS) in the European subset of the CFSRS where genotype data, as well as clinical group data (no disorder, SSD only, language impairment (LI) only, SSD+LI, CAS) were available. Risk scores were derived from association statistics from our CFSRS GWASs (see GWAS methods section for details) and were constructed using PLINK 1.9^46^ (clump and score functions). Additional details are in the Supplemental Methods. These polygenic risk scores were used to examine the hypothesis that an increase in PRS score would associate with more complex clinical phenotypes when comparing SSD only versus SSD+LI and CAS.

### Statistical analysis of Methylome-wide data

#### Methylome-wide association study (MWAS)

Quality control analysis of methylation data is detailed in Supplemental Methods. We tested for association between CpG beta values and endophenotypes using the linear mixed model approach of GRAMMAR-Gamma^45^ as implemented in RVtests^44^. Because our phenotypes were age-normed, we did not adjust for age, but rather for sex and one to four PCs. We also examined methylation-QTLs (meQTL) as described in the Supplemental Methods.

### Replication dataset – ALSPAC

To replicate our GWAS findings, we obtained data from the Avon Longitudinal Study of Parents and Children (ALSPAC). The ALSPAC study was a prospective population-based birth cohort of babies born from > 14,000 pregnancies between April 1991-December 1992, who were followed prospectively with a wide battery of developmental tests, parental questionnaires, child-completed questionnaires, and health outcomes^47-49^. Pregnant women resident in Avon, UK with expected dates of delivery 1st April 1991 to 31st December 1992 were invited to take part in the study. The study website contains details of all the data that is available through a fully searchable data dictionary (http://www.bris.ac.uk/alspac/researchers/data-access/data-dictionary). Ethical approval for the study was obtained from the ALSPAC Ethics and Law Committee and Institutional Review Board of Case Medical Center and University Hospitals. Because this was a birth cohort, all children were included, regardless of diagnosis. We obtained both parental report data on speech development in the children, and also communication measures similar to those that we analyzed (see Communication Measures above and Supplemental Table 3). As this was a longitudinal study, different measures were given at different ages, and when the same domain was tested at two different ages, the identical measure was not used. At some ages, only random subsets were selected, so the sample size available from each age is not the same. In Supplemental Table 4, we list the measures given in the CFSRS battery along with the most similar measure given in ALSPAC.

#### GWAS in ALSPAC data

QC analyses of ALSPAC data are described in Supplemental Methods. Because of the format of data that were provided, we used slightly different methods for statistical analyses. Genetic association testing was performed using linear regression in Hail 0.1. Covariates adjustments included sex and the first two PCs. Age was not a consideration as ALSPAC is a longitudinal birth cohort study and age differences were negligible for any given measure.

### Functional annotation and results integration

In this analysis, we considered CFSRS the discovery sample, since families were ascertained through a child with SSD, and used ALSPAC as the replication sample. We identified associated loci with SNPs significant at p<10^−5^ in CFSRS and p<0.05 in ALSPAC, with effects in the same direction.

#### Functional annotation

Because the majority of our findings are intergenic and/or fall in noncoding regions, we relied on annotation tools FUMA and HaploReg to characterize which genes our variants might affect, as well as variants’ functionality. We utilized FUMA^50^ for mapping genes to our variants based on genomic proximity, eQTL evidence and chromatin interactions evidence. Default settings in FUMA were used, with the exception of tissue specificity. We hypothesized that gene expression and regulation would be most relevant in brain and neural tissues, as well as muscles related to speech. In FUMA we focused on eQTL and chromatin interaction evidence in our target tissues (brain, muscle and esophagus). HaploReg v.4.1 was used to examine the chromatin state evidence predicting whether the variant fell in a promoter or enhancer region. In HaploReg we focused on chromatin state evidence in our target tissues (brain and muscle).

#### Locus prioritization

In order to further prioritize and synthesize our findings, we annotated associated loci as described above, including annotation of associated effects of these loci in the literature, and incorporated supportive findings from our MWAS. We summarize findings in Table 2, and generated a simple locus priority score as the number of times a locus included an enhancer and/or promoter, included an eQTL, was previously associated with a communication disorder and/or neuropsychiatric disorder, showed eQTL or chromatin state evidence specific to brain and/or neural tissues, mapped to a gene that was a *FOXP2* target in brain tissue ^51-53^, and a meQTL in that region (at p< 5×10^−5^) with an associated methylation site (at p< 0.05) with the same phenotype as the associated GWAS loci. We applied the EpiXcan pipeline^54^ to identify eQTLs with our associated SNPs that are differentailly expressed in the dorsolateral prefrontal cortex (DLPFC)^55^ (Supplemental Methods).

**Table 2.**
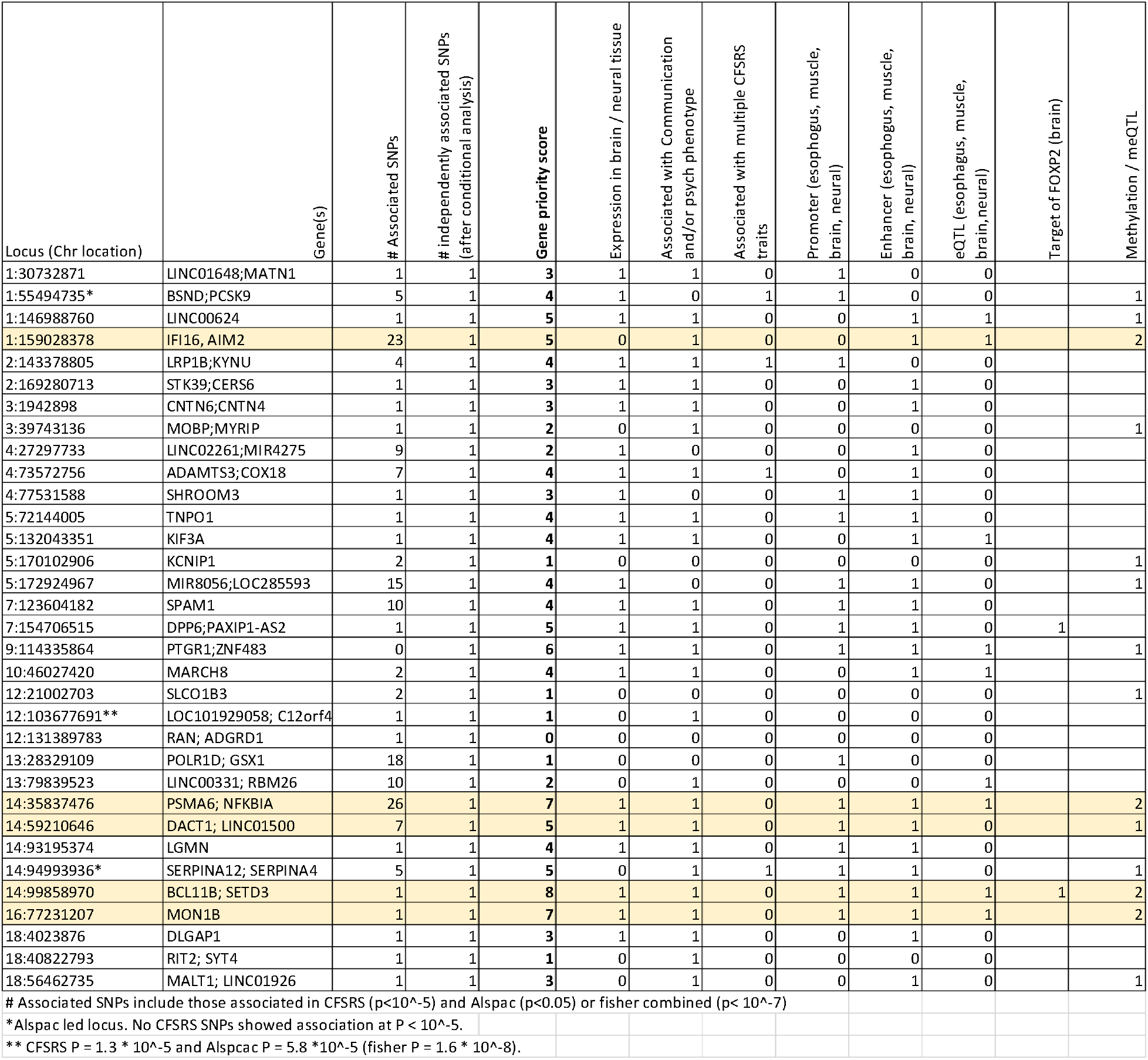
Annotation of most significant loci with replication in CFSRS and ALSPAC

## RESULTS

The CFSRS sample included 435 subjects from 148 families (Table 1). Of these, 27% had SSD only, 4% had LI only, 16% had SSD+LI without CAS, and 11% had CAS (Table 1). Of the subjects in the ALSPAC sample, the prevalence of speech problems by parental report varied from 4%-6% (Supplemental Table 3).

### Genetic correlation analysis reveals new relationships among endophenotypes

Genetic correlation analysis revealed that while many of the patterns of correlation were consistent with phenotypic correlations we have previously reported^10^, polygenic correlations enable a deeper understanding of these measures, which will inform examination of replication of association effects both within the CFSRS data set and with measures from ALSPAC (Figure 1). For example, while previous studies have demonstrated a strong genetic correlation between reading and spelling measures, polygenic correlation analysis additionally reveals correlations between those skills and Elision. Not surprisingly, expressive and receptive language, as measured on the CELF, are strongly correlated with vocabulary (EOWPVT and PPVT) in addition to reading (WRMT-AT and WRMT-ID). Vocabulary is also strongly correlated with listening comprehension (WIAT-LC).

**Figure 1.**
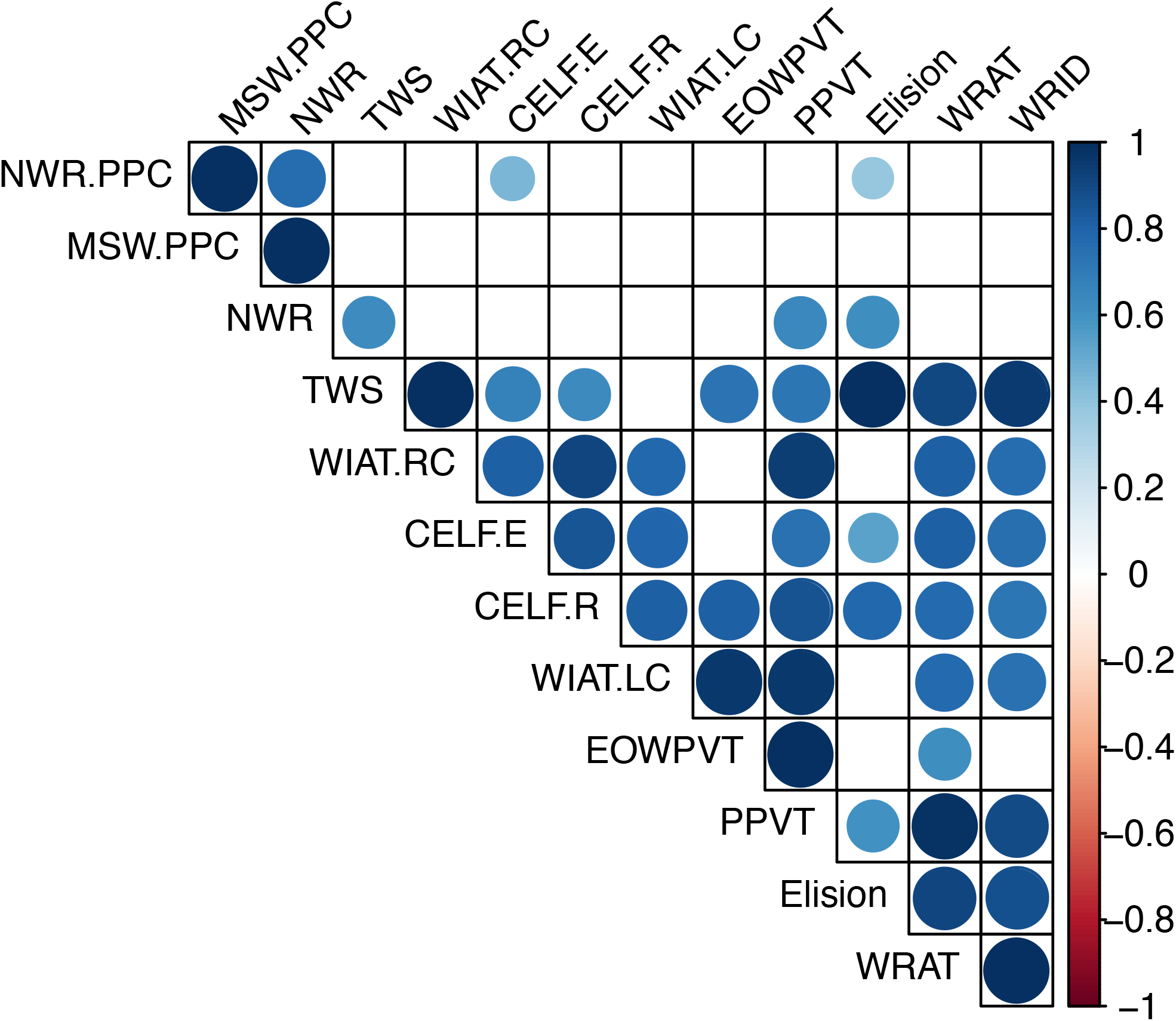
Genetic correlation matrix across traits in CFSRS. Figure 1 shows cross-trait correlation results for each pair of tests using GCTA’s bivariate REML analysis. Cross-trait correlation was tested under the null hypothesis of 0 correlation. Circles shown are for results significant at *P*<0.05, with increasing diameter/color corresponding with increasing correlation (circles omitted otherwise).

### Most significant findings from GWAS reveal 5 new candidate genes

The majority of associated SNPs (p<10^−5^) were intergenic, with a lesser number of intronic SNPs (Supplemental Figure 2). Noncoding regions harboring a significant proportion of risk alleles is consistent with previous findings related to neuropsychiatric disease and behavioral traits^56^. We focused on SNPs that had a p-value<1×10^−5^ in CFSRS with replication with a related trait in ALSPAC (p<0.05), or Fisher combined p-values < 1×10^−7^, that had functional relevance based on our gene priority score (Table 2).

**Figure 2.**
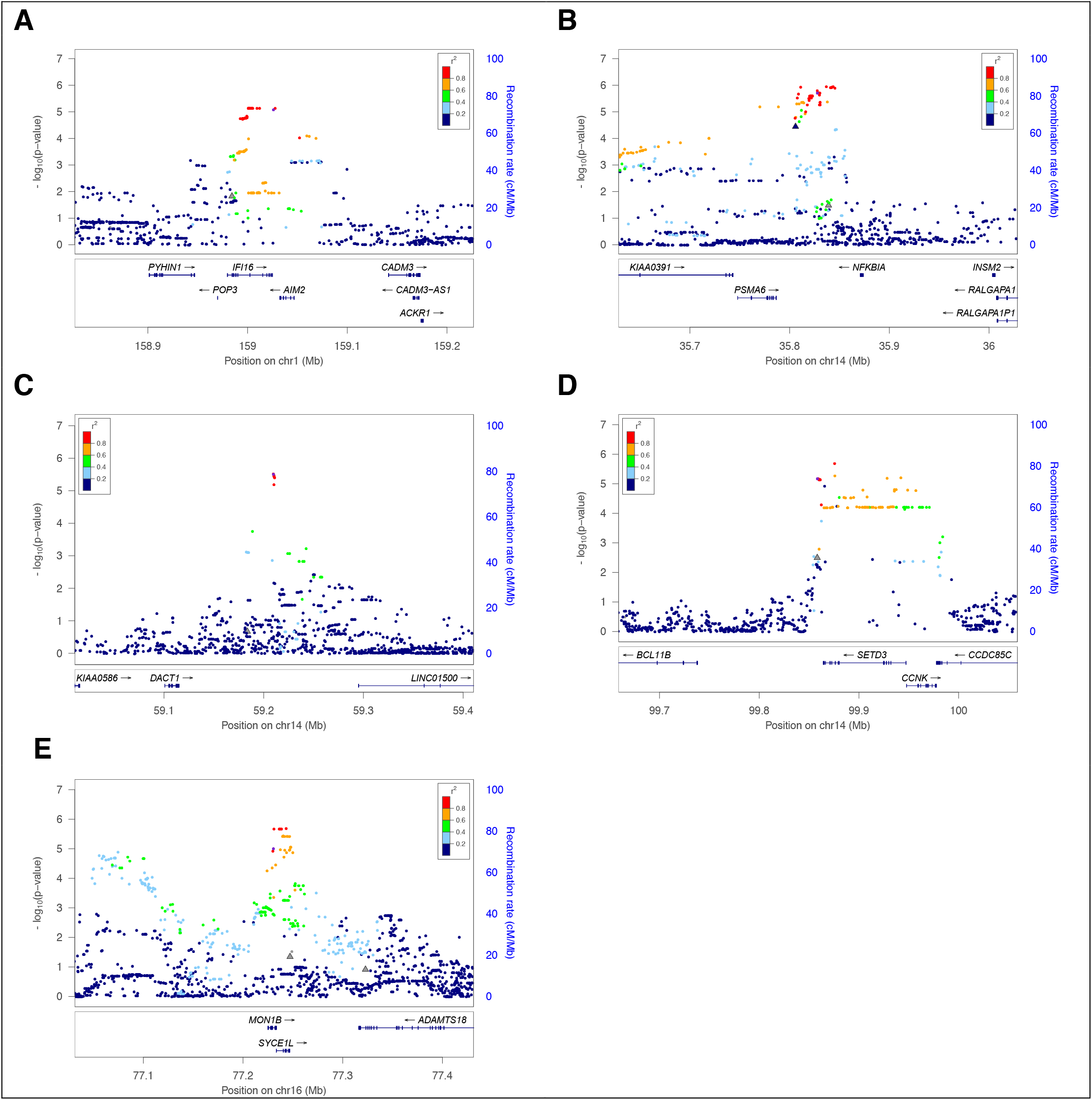
Locus zoom plots for most signfiicant findings. Figure 2 shows association results for the top loci. P-values displayed are for CFSRS and are for the test for which the top SNP was observed. Circles show P-values for SNP associations and triangles show P-values for methylation associations (specifically those for which the top SNP is an meQTL). The larger plot shows the top SNP for each region +/-200 kb. The window highlights the region that spans significant association results (P≤1×10^−5^ in CFSRS. **A**. IFI16 region (window spans chr1:159001292-159028378) rs855865 was associated with NSW in CFSRS (p=7× 10^−6^) and with vocabulary (WISC-V) in ALSPAC (p=0.01). This region also includes an meQTL (rs12124059, p=4×10^−8^) for methylation marker cg07196514, and this methylation marker was also associated with NSW (p=0.018). **B**. NFKBIA region (window spans chr14:35770806-35846092). rs57645874 was associated with Elision in CFSRS (p=1 × 10^−6^) and with reading accuracy (NARA-A) in ALSPAC (p=0.02). This region also contains an meQTL, rs4981288, for cg07166546 (p=2×10^−50^), and this methylation marker was associated with Elision (p=3. ×10^−5^), TWS (p= 0.0005) and WRMT-ID (p=0.002). **C**. DACT1 region (window spans chr14:59210335-59221002). rs856379 was associated with MSW in CFSRS (p=3×10^−6^) and with nonword reading (ALSPACread) in ALSPAC (p=0.036). This SNP is an meQTL for methylation marker cg13972423 (p=3×10^−5^), **D**. SETD3 region (window spans chr14:99858970-99942692). rs1257267 was associated with WRMT-AT in CFSRS (p=6.58×10^−6^) and with nonsense word repetition (CNrep5) in ALSPAC (p=0.05). While only 1 SNP replicated between CFSRS and ALSPAC, 14 additional SNPs showed association in CFSRS at p<10^−5^. This SNP is an meQTL for cg18949721 (p=4×10^−12^), which was also associated with WRMT-AT (p=0.003). **E**. MON1B region (window spans chr16:77231207-77248555). rs4888606 was associated with MSW in CFSRS (p=9 × 10^−6^) and with nonword reading (ALSPACread) in ALSPAC (p=0.046). While only 1 SNP replicated between CFSRS and ALSPAC, 18 additional SNPs showed association in CFSRS at p<10^−5^. This SNP falls in an intron of *MON1B* and is an meQTL for cg06128999 (p=4×10^−23^) and cg05007098 (p=1×10^−15^), which were also associated with MSW (p=0.045 and p=0.12, respectively). Functional annotation is in Supplemental Figure 2.

Among the 5 prominent loci, all had enhancers or promoters for muscle, brain, and/or neuronal progenitor cells, 4 out of 5 had significant methylation and meQTL effects, and 3 out of 5 had eQTLs for brain and/or skeletal-muscle tissue (Figure 2, Supplemental Table 5). EpiXcan analysis suggested that the SNP in the chromosome 1 *IFI6* region is associated with expression in the DLPF cortex (Elision p=0.018, TWS p=0.008; Supplemental Tables 6 and 7). The first region on chromosome 14, including *NFKBIA* and *PPP2R3C*, shows significant chromatin interaction mapping in adult cortex tissue. *NFKBIA*, which codes for a component of the NF-κB pathway, is associated with neurogenesis, neuritogenesis, synaptic plasticity, learning and memory^57^. The second region on chromosome 14 includes *PP2R3C*, which is within the topologically associating domain (TAD) boundary of the *NFKBIA* locus in Hippocampus and DLPFC. EpiXcan analysis showed *NFKBIZ*, a gene in the same pathway as *NFKIBA*, is also associated with expression in the DLPFC (Elision p=0.000452, TWS p=0.004939; Supplemental Tables 6 and 7).

### Replication of previous communication disorder loci

*ATP2C2* was associated with WRMT-ID (p=7.6×10^−8^), WRMT-AT (p=4.6×10^−5^), and Elision (p=4.6×10^−5^), consistent with prior literature^58^ (Supplemental Figures 3 and 4). Similarly, *CYP19A1* was associated with WRMT-AT (p=2.8×10^−5^), Elision (p=3.3×10^−4^), and WRMT-ID (p=5.0×10^−4^), validating a previous association^59^. *CNTNAP2* was associated with CELF-R (p=5.2×10^−6^), and DDK (p=2.9×10^−5^), replicating a previous association^58^. While SNPs within *ROBO1* and *ROBO2* were not significantly associated with our measures, SNPs in the intergenic region were associated with WRMT-ID (p=3.6×10^−6^); *ROBO1* was originally associated with dyslexia while *ROBO2* was originally associated with expressive vocabulary^20,60^. Finally, SNPs within the *DCDC2-KIAA0319-TTRAP* and in *FOXP2* regions were associated with various traits at p<0.01. Within the ALSPAC cohort, a different pattern of replication emerged (Supplemental Figure 5), with sometimes different SNPs and/or different phenotypes than those associated with CFSRS.

**Figure 3.**
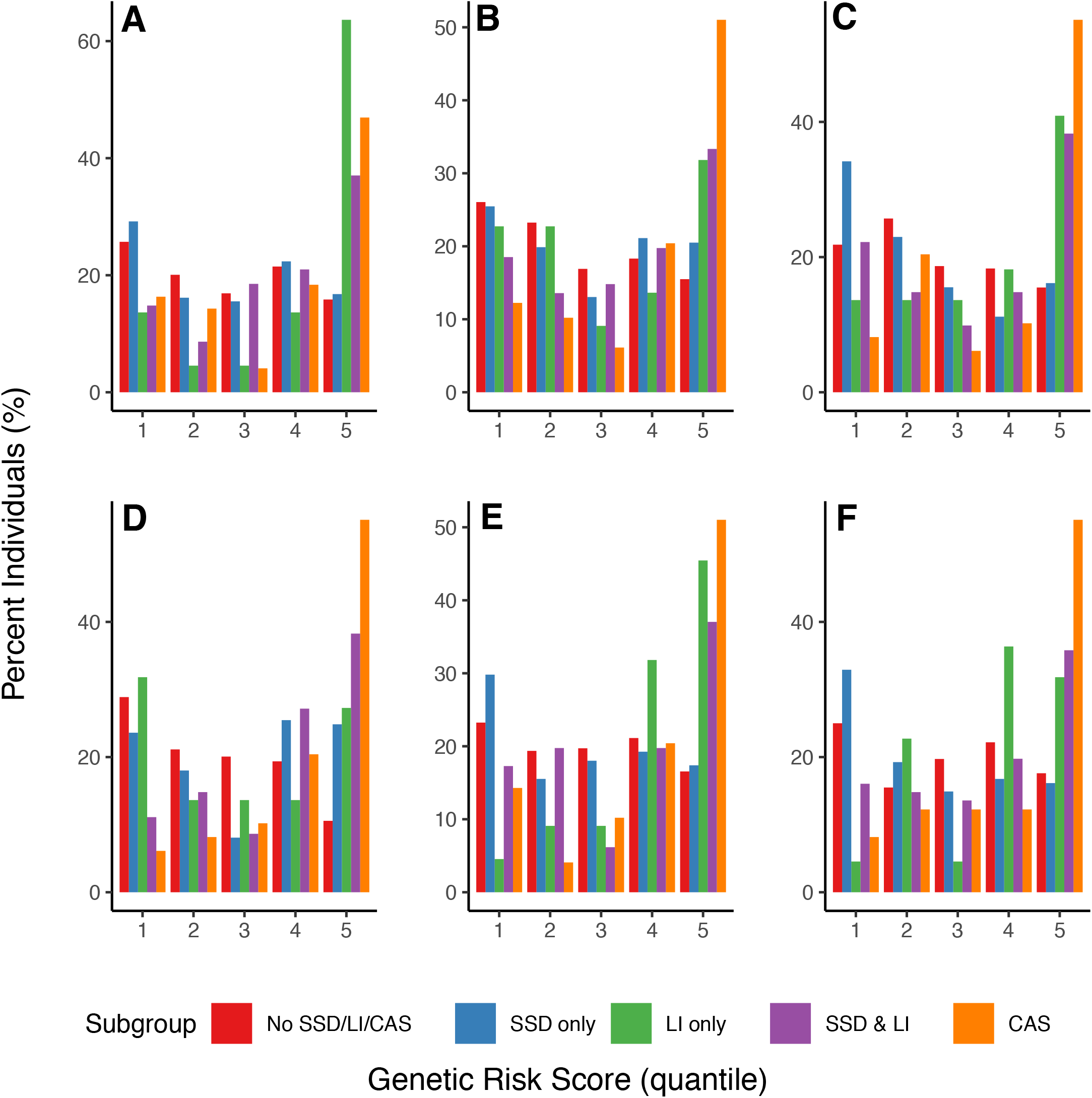
Polygenic risk scores across major domains. We constructed polygenic risk scores for 587 individuals who were both genotyped and had clinical subgroup information available. Polygenic risk scores are displayed by quantile across the clinical subgroups for six endophenotypes representing the major domains (A Receptive language; B Expressive vocabulary; C Phonological awareness; D Phonological memory; E Spelling; F Reading decoding).

In addition, we examined loci (genes and/or SNPs) associated in recently published GWAS studies of language and reading^13-20^(Supplemental Table 8); we restricted our examination to the CFSRS data, since the ALSPAC data were included in some of the published studies. In these analyses, we often observed cross-trait replication, with most genes originally associated with dyslexia, and associated with other traits in our sample. These included *ZNF385D*^14^, which was associated with all CFSRS traits at p<0.005, *CDH13*^19^, associated with all CFSRS traits at p<0.005, *GRIN2B*^15^, associated with TWS, EOWPVT, and Elision at P<0.0005 and all CFSRS traits at P<0.05, *NKAIN*^15^, associated with CELF-R at 9.7 × 10^−5^ (rs16928927 p=1×10^−4^) and WIAT-RC (p=4×10^−4^), and *MACROD2* ^17^ associated with all CFSRS traits at p<0.005).

### Polygenic risk scores are associated with increasing clinical severity

In Figure 3, we illustrate polygenic risk scores (PRS) for 6 endophenotypes representing the major domains (receptive language, expressive vocabulary, phonological awareness, phonological memory, spelling, and reading decoding), by quintile, across the clinical subgroups (all endophenotypes are illustrated in Supplemental Figure 6). Generally, we found that polygenic load, indicated by increasing risk scores, was associated with clinical severity (p<1×10^−8^ by ANOVA), with typical children having the lowest scores, followed by children with SSD-only, and children with SSD+LI and CAS having the greatest scores. The exception to this trend is receptive language, where the genetic load is greatest for children with LI, for whom receptive language is a focal deficit. Thus, in general, an increase in PRS score is associated with greater clinical severity.

## DISCUSSION

Communication disorders are genetically complex, manifested by a variety of deficiencies in articulation, vocabulary, receptive and expressive language, phonological awareness, reading decoding and comprehension, and spelling. This GWAS ascertained children through an earlier-presenting clinical disorder and examined several key communication measures, and is thus one of the first studies of its kind. This study is also novel in that it is the first GWAS to include a measure of phonological awareness, as well as a motor speech measure. By analyzing several endophenotypes together, we can draw conclusions about the common genetic basis across these seemingly dissimilar skills. Here, we have identified five new candidate regions, some containing multiple genes, that have connections to neurological function and regulation of neurological pathways. We also found that increased polygenic load is associated with more severe communication disorders. Finally, by examining genetic correlations among these traits, we conclude that different domains of communication have some common genetic influences. All of these aspects together add new clarity regarding the genetic underpinnings of speech and language skills.

First, the novel candidate genes that we have identified all have roles in neurological function as evidenced by expression levels of those genes in brain and/or neural tissue, and associations with other communication and/or psychiatric phenotypes. This commonality between communication traits and brain and neural pathways was also demonstrated by a mouse study of vocalization^61^, and pleiotropy between brain, learning, and psychiatric phenotypes was recently demonstrated by a large GWAS of brain phenotypes^62^. Existence of enhancers, promoters, and methylation effects in the associated regions further emphasizes the importance of regulatory effects on these traits. Deletions spanning *SETD3* and *CCNK* have been associated with syndromic neurodevelopmental disorders^63^ and variants in *SETX*, within this same family of genes, have been associated with CAS^64^. In addition, *CCNK* is in the *FOXP2* pathway in brain tissue^51-53^. *NFKBIA* is involved in regulation of the NF-κB pathway, which is involved a number of brain-related processes including neurogenesis, neuritogensis, synaptic plasticity, learning, and memory^65^. *PPP2R3C* has been associated with schizophrenia^66^. *IFI6* expression has been associated with autism^67^ and overexpression of *IFI6* in the brain is present in chronic neurodegeneration^68^. Finally, *DACT1* may be involved in excitatory synapse organization and dendrite formation during neuronal differentiation^69^ and is mainly expressed within the first two trimesters of pregnancy, just before the first evidence of speech processing is observed in preterm neonates^70^. Interestingly, *SETD3, NFKBIA*, and *IFI6* are all also tied to the immune system, and a recent study identified an excess of T cells in brains of individuals with autism^71^.

Second, understanding the genetic architecture across these endophenotypes is essential for understanding how loci are associated with different measures in different study cohorts or across the developmental trajectory. Strong genetic correlations are observed between spelling, reading comprehension and decoding, expressive and receptive language, vocabulary, and phonological awareness. The strongest replications were for a variety of measures collected in CFSRS with ALSPAC from older youth. Consistent with these findings, we previously demonstrated that spelling at later ages has a higher estimated heritability than spelling at school-age^11^. Measures administered in older youth may also be more sensitive to variations in clinical manifestation of SSD. Examination of the ALSPAC measures suggests that many of those administered at younger ages may have tapped different domains than intended, or may have been less sensitive to later emerging reading and spelling skills. Methods of cohort ascertainment may also be important in comparing our findings to those of other studies. Our families were ascertained through a child with SSD whereas other studies ascertained subjects through LI or dyslexia. These different ascertainment schemes affect both the available measures, as well as the distribution of scores and power to detect association. Since both LI and dyslexia emerge later than SSD, longitudinal studies that ascertain through a proband with SSD will be able to capture variants associated with all three disorders, as there is high comorbidity. In addition to the plethora of studies ascertaining children at a variety of ages, which has an impact on the heritability of traits^10^, these studies use a wide variety of measures, even for the same endophenotype. Moreover, these studies have been conducted in populations that speak different languages of varying orthographic transparency, which makes them difficult to compare. As noted by Carrion-Castillo et al.^13^, most of the novel loci identified through GWAS have been unique to each study, and these aforementioned issues may explain that lack of replication. Thus, examination of the genetic correlation matrix is essential for interpretation of results across studies, as it is nearly impossible to analyze the same exact traits, as we have demonstrated with our replication study cohort (ALSPAC).

Third, we replicated candidate genes that had been previously primarily associated with reading and/or language impairment: *CNTNAP2, ATP2C2*, and *CYP19A1*. These analyses extend previous findings to show that these genes are associated with articulation (*CNTNAP2*) and phonological awareness (*ATP2C2* and *CYP19A1*). This further illustrates the pleiotropic nature of these genes. While we did not observe association with SNPs within the coding regions of *ROBO1* and *ROBO2*, we did observe significant associations with SNPs between these two genes, which may have regulatory influences on *ROBO1/ROBO2*. We also replicated (p<5×10^−3^) loci identified in recent GWAS of reading and/or language traits. Similar to another association study between *FOXP2* variants and language^72^, we did not observe statistically significant association between *FOXP2* and measures in CFSRS, though there was replication of some traits at a less stringent (p<0.01) level^72^.

Finally, our analysis of polygenic risk scores shows strong associations between these risk scores and clinical outcomes of increasing severity. Because of the strong significance of these findings, this suggests that the genetic architecture of communication disorders maybe largely polygenic, which may additionally explain the lack of replication and/or genome-wide significance. While other studies have examined polygenic risk scores associated with language^15,73^, ours is the first to examine polygenic risk associated with other communication endophenotypes. It is noteworthy that our associated SNPs fell outside of gene coding regions but resided in regulatory regions, even having potential regulatory effects themselves. This further illustrates the genetic complexity of communication disorders; perhaps the search for single gene dysfunction is misplaced, and rather regulatory functions are more relevant.

This study has several limitations. The sample size of the CFSRS cohort was modest, potentially reducing power. There was not clear correspondence between measures obtained in ALSPAC with those in CFSRS, necessitating consideration of cross-trait replication. We restricted analyses in both cohorts to individuals of European descent because of low sample size in other ethnic groups, reducing generalizability.

In summary, this first GWAS of communication measures ascertained through families with SSD identified five new candidate genes, all with potential relevance in central nervous system function. Polygenic risk is strongly associated with more severe speech and language outcomes. Careful consideration of genetic correlation among domains of verbal and written language shows that these loci have general effects on communication, not specific to any single domain, suggesting a common genetic architecture. Further research is needed to more closely examine the impact of regulatory variants on these outcomes.

## Data Availability

Individual-level data from the Cleveland study are not available for broad sharing because of IRB restrictions; summary statistics may be obtained by request from Dr. Sudha Iyengar.
ALSPAC data are available through an application process to ALSPAC.

## ACKNOWLEDGMENTS

We would like to thank the families who have so generously participated in this study for many years. This research was supported by the Genomics Core Facility of the CWRU School of Medicine’s Genetics and Genome Sciences Department. This work made use of the High Performance Computing Resource in the Core Facility for Advanced Research Computing at Case Western Reserve University. This work was supported by NIH grant R01DC000528 awarded to Dr. Lewis and R01DC012380 awarded to Dr. Iyengar. We are extremely grateful to all the families who took part in the ALSPAC study, the midwives for their help in recruiting them, and the whole ALSPAC team, which includes interviewers, computer and laboratory technicians, clerical workers, research scientists, volunteers, managers, receptionists and nurses. The UK Medical Research Council and Wellcome (Grant ref: 217065/Z/19/Z) and the University of Bristol provide core support for ALSPAC. This publication is the work of the authors and Dr. Sudha Iyengar will serve as guarantor for the contents of this paper. GWAS data for ALSPAC was generated at the Genotyping Facilities at Wellcome Sanger Institute.

## DATA AVALABILITY

Data from the Cleveland Family Speech and Reading study are not available for broad genetic data sharing because of IRB restrictions. Please contact the corresponding author, Dr. Sudha Iyengar, to request data, which will require an IRB application.

## MATERIALS AND CORRESPONDANCE

Please contact Dr. Sudha Iyengar, regarding access to summary statistics.

## DESCRPTION OF SUPPLEMENTAL DATA

Supplemental Methods: Describes behavioral phenotypes in detail and detailed methods for genetic methylation analysis

Supplemental Tables and Figures:

Supplemental Table 1. Descriptive statistics for CFSRS measures

Supplemental Table 2. Results of methylation analysis of candidate gene regions

Supplemental Table 3. Descriptive statistics for ALSPAC sample

Supplemental Table 4. Correspondence between CFSRS and ALSPAC measures

Supplemental Table 5. Annotation of functional implications of most significant loci from GWAS

Supplemental Table 6. PsychEncode EpiXcan method using Meta-analysis results of Elision GWAS

Supplemental Table 7. PsychEncode EpiXcan method using Meta-analysis results of TWS GWAS

Supplemental Table 6 – Association results from regions identified from published GWAS of reading and language phenotypes

Supplemental Figure 1. Distribution of associated SNPs

Supplemental Figure 2. Functional annotation corresponding to Figure 3.

Supplemental Figure 3. Clustering of Significant Variants (P < 0.01) among Known Speech Genes across CFSRS Tests

Supplemental Figure 4. LocusZoom plots of candidate genes where at least one trait had a SNP significant at p < 10^−4^

Supplemental Figure 5. Clustering of Significant Variants (P < 0.01) among Known Speech Genes across ALSPAC Tests

Supplemental Figure 6. Polygenic Risk score across all individual measures

---

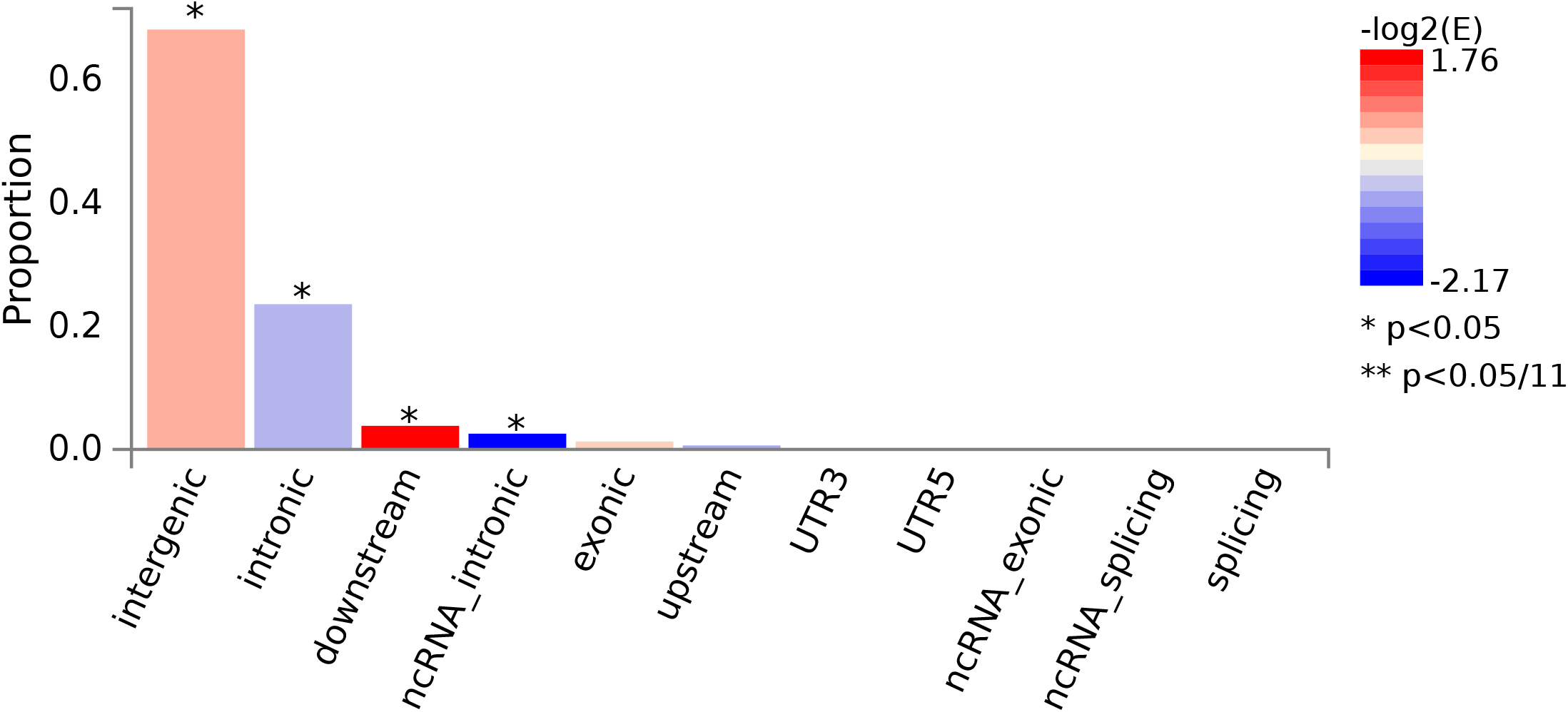

