## Supplemental Methods, Tables, and Figures for "Association between genes regulating neural pathways for quantitative traits of speech and language disorders"

**Subject ascertainment – inclusion criteria**

All participants met inclusion criteria based on information provided by a parent in an interview or via questionnaire including: normal hearing acuity; fewer than six episodes of otitis media prior to age 3; monolingual English speaker; absence of a history of neurological disorders other than childhood apraxia of speech (CAS), such as cerebral palsy or autism spectrum disorder; and a diagnosis of a SSD or suspected CAS by a local speech-language pathologist or neurologist.

**Measures – Cleveland Family Speech and Reading Study (CFSRS)**

As these measures were collected as part of a longitudinal study, the first available assessment was used for analysis.

*Oral Motor Skills Test*

The *Robbins and Klee Oral Speech Motor Control Protocol* ^1^ (DDK-OSMCP) assessed oral motor skills in children 4 to 6 years. Diadochokinetic rates on single, double and multi-syllables were timed and scored. The *Fletcher Time-by-Count Test of Diadochokinetic Syllable Rate ^2^* (DDK-FL) was administered to children 7 years or older. This test also assesses rates of syllable repetition on single, double and multi-syllable sequences. Z-scores were generated, and the scores for single and multi-syllables were combined for analysis. A shorter time for syllable sequence repetition indicates better oral motor skills. Because the DDK-OSMCP is scored in the opposite direction, these values were negated prior to merging with the DDK-FL; for this merged score, lower scores imply worse performance. We shall refer to this merged variable as DDK.

*Expressive and Receptive Vocabulary*

The *Expressive One Word Picture Vocabulary Test-Revised* (*EOWPVT ^3^)* assesses expressive vocabulary and requires the examinee to name pictures representing objects, actions, and concepts. The *Peabody Picture Vocabulary Test- Third Edition (PPVT-III ^4^)* tests receptive vocabulary and requires the examinee to point to the image that best matches the stimulus from a group of four*.* Both of these measures are given to children ages 2 years or older.

*Multisyllabic Nonword and Real Word Repetition*

*Nonsense word repetition (NWR ^5^)* requires children to repeat 15 non-words, a task which requires encoding of unfamiliar phonological sequences; deficits in encoding would result in inaccurate word repetition^5^ and can discriminate children and adults with resolved SSD from those who never had SSD ^6^. While this task is normally given to individuals ages 4 years through adulthood, the children with CAS generally could not perform this task in preschool, so the first available assessment starting at age 7 was used for this analysis. The Multisyllabic Word Repetition task (MSW) requires children to accurately sequence phonemes by repeating real multisyllabic words. Target words include aluminum, thermometer, and sympathize ^5^. The test is scored by determining the percentage of words repeated correctly. In addition, we scored the percentage of phonemes repeated correctly; these tests are the MSW-PPC and NSW-PPC.

*Phonological Awareness Measure*

The *Elision* subtest of the *Comprehensive Test of Phonological Processing* ^7^ requires the individual to repeat a word and then say the word with a sound or syllable deleted. The Elision subtest is a measure of phonological awareness, that is knowledge of the sound system of oral language. Children with poor phonological awareness skills demonstrate weakness in single word reading. In addition, we gave an earlier version of the Elision task, Comprehensive Test of Phonological Processing-Experimental Version (Wagner, Torgesen, & Rashotte, 1994, personal communication), which we combined with this CTOPP subtest in order to create a single variable for analysis. We transformed the CTOPP Elision subtest into a z-score prior to merging with the original Elision.

*Rapid Automatized Naming*

*The Rapid Color Naming (RAN)* requires the individual to name colored squares as quickly as possible. The total number of seconds to name all the colors is the individuals score. This test measures the subject’s ability to retrieve phonological information from long-term memory and is predictive of reading decoding skills. We had an older version of this task^8^ as well as the CTOPP subtest. We transformed the CTOPP into a z-score and flipped (or negated) the RAN prior to merging CTOPP colors, a scaled score with a mean of 10 and standard deviation of 3, and RAN colors, a z-score with larger values representing worse performance.

*Spoken Language Measures*

There were two language assessments that were combined, Clinical Evaluation of Language Fundamentals- Revised (CELF-R) ^9^ and the Test of Language Development- Primary- Second Edition (TOLD-P2) ^10^. Participants enrolled in the study after the release of a newer version of the tests were given the new version. Both the CELF-R and TOLD-P2 provide standard scores for receptive and expressive language skills. We merged the CELF-R receptive with the TOLD-P2 listening quotient, with preference to the CELF-R if both were given at the same time point. Both the CELF-R and the TOLD-P2 were scaled scores, with a population mean of 100 and standard deviation of 15. We merged the CELF-R expressive with the TOLD-P2 speaking quotient, with preference to the CELF-R if both were given at the same time point. Both the CELF-R and the TOLD-P2 were scaled scores, with a population mean of 100 and standard deviation of 15.

*Reading Measures*

The *Woodcock Reading Mastery Test-Revised, Word Attack subtest (WRMT-AT ^11^*) evaluates phonetic decoding skills by requiring examinees to read a list of 45 non-words; the test includes nonwords such as *ip, din, ceisminadolt, and gnouthe*. The *Woodcock Reading Mastery Test- Revised, Word Identification Subtest (WRMT-ID ^11^)* assesses single word reading ability by requiring examinees to read a list of 106 real words. This task is given to individuals ages 5 through adulthood. The *Wechsler Individual Achievement Test* ^12^ has 2 subtests. The *Reading Comprehension subtest* (WIAT-RC) consists of printed passages that the individual reads and then answers orally presented question on the passage. It assesses the individual’s ability to recognize details and make inferences concerning what has been read. The reading comprehension subtests provides information on the child’s ability to use picture cues, recognize detail, sequence events, identify cause-effect relationships, make inferences, and compare and contrast characters, objects or events from the passage. The *Listening Comprehension* subtest (WIAT-LC) consists of orally presented passages that are sometimes paired with pictures. The child understanding of the passage is examined by answering questions about the passage. The Listening Comprehension subtest, similar to the Reading Comprehension Subtest, assesses the child’s ability to use picture cues, recognize detail, sequence events, identify cause-effect relationships, make inferences, and compare and contrast characters, objects or events from the passage.

**Age adjustment in CFSRS data**

For the following tests - NSW, NSW PPC, MSW and MSW PPC - we did not have population normed data, therefore, we converted all scores to age-adjusted z-scores using CFSRS controls. Here, controls were defined as individuals without SSD, LI or CAS. To age-adjust we chose the first available observation for each of the four tests for every control within the CFSRS to determine the effect of age. The age-adjusted score is simply the standardized residual of the score with the effect of age and age-squared regressed out (where the age effect is determined by controls and subsequent adjustment is applied to all participants)^13,14^. Age and age-squared are both used to determine the effect of age, as there is a non-linear relationship between age and each of the above four tests. If applicable, test scores were transformed to an approximately normal distribution using the Box-Cox power transformation^39^. Because measures were already age-normed or age adjusted, age was not included additionally as a covariate in GWAS or other analytical models.

**ALSPAC Study**

*Ascertainment*

The ALSPAC study was a prospective population-based birth cohort of babies born from > 14,000 pregnancies between April 1991-December 1992, who were followed prospectively with a wide battery of developmental tests, parental questionnaires, child-completed questionnaires, and health outcomes^47-49^. Pregnant women resident in Avon, UK with expected dates of delivery 1st April 1991 to 31st December 1992 were invited to take part in the study. The initial number of pregnancies enrolled is 14,541 (for these at least one questionnaire has been returned or a “Children in Focus” clinic had been attended by 19/07/99). Of these initial pregnancies, there was a total of 14,676 fetuses, resulting in 14,062 live births and 13,988 children who were alive at 1 year of age. The study website contains details of all the data that is available through a fully searchable data dictionary

(<http://www.bris.ac.uk/alspac/researchers/data-access/data-dictionary>). Ethical approval for the study was obtained from the ALSPAC Ethics and Law Committee and Institutional Review Board of Case Medical Center and University Hospitals. Blood samples were also collected for biomarker and genetic analyses.

*Communication Measures*

We worked with the ALSPAC team to find the most equivalent measures to match those in the CFSRS. These are summarized in Supplemental Table 4. The *multisyllabic word repetition (MWR)* task consisted of repeating “buttercup” and dinosaur” 5 times each – the total correct score was used for analysis. This task was given at age 5. A *nonsense word repetition task ^15^ (CNrep)* was given at age 5 (*CNrep5*), and a shortened version of this task was given at age 8 (*CNrep8)*. Two reading tasks were given. At age 7, both the *Wechsler Objective Reading Dimensions* *single word reading task (WORD)* and *Neale analysis of reading ability (NARA)* were given. The NARA has a reading comprehension (*NARA-C*) and reading accuracy (*NARA-A*) subtest. A non-word reading task was designed specifically for ALSPAC, so we refer to it as *ALSPACread*. The spelling tasks were based on work by Nunes et al.^16^, so we refer to this task as *ALSPACspell7* and *ALSPACspell9* for the tests given at ages 7 and 9, respectively. The *Wechsler Objective Language Dimensions* (*WOLD)* is a test of expressive language ability in children, and has several subtests that were used for language and vocabulary assessment: *comprehension (WOLD-C*), *expression (WOLD-E)*, and *vocabulary (WOLD-V)*; these were all given at age 8.

Because all the children were the same age when specific assessments were given, no age adjustment was needed. There were no equivalent measures for RAN and Elision.

**Molecular methods**

DNA was extracted from buffy coats or saliva samples as previously described^9^. All genotyping was performed using the Illumina Omni 2.5 platform. Standard QC procedures were applied, including filtering based on call rate, Hardy-Weinberg equilibrium (HWE), chromosome (autosomes only), minor allele frequency (MAF) and Mendelian errors. Principal components analysis was conducted using markers that attained MAF ≥ 0.01, sample and variant call rate ≥ 0.98 and p≥0.0001 from an exact test of HWE, while omitting genomic regions with long range linkage disequilibrium (LD)^17^. Genotyped data were later imputed to the Phase 3, cosmopolitan reference option, of the 1000 Genomes Project panel using the University of Michigan Imputation server^18^ which implements minimac3 ^19^. Following imputation, all markers with imputation quality score R^2^<0.6 and MAF<0.05 in our population were removed. Samples were processed and typed for the Illumina Methylation450 chip by the CWRU School of Medicine Genomics Core. The final data set typed for the Methylation450 panel comprised 713 unique individuals, plus 60 duplicate samples.

**Methylome-wide association analysis (MWAS)**

*Methylation QC*

Quality control and normalization of raw methylation data (as Illumina .idat files) were carried out using the Bioconductor package RnBeads for R^20^. We removed methylation probes in non-CpG contexts, with nearby SNPs, on the X and Y chromosomes, and probes with low variability (SD < 0.005), leaving a total of 470,870 CpG markers with detection p value < 0.05. We normalized signal intensity by means of the BMIQ algorithm ^21^, which adjusts for differences between Infinium I and II loci, and adjusted background by the methylumi NOOB procedure, as implemented in RnBeads. Our final data set was scaled to proportion of methylated DNA strand (β) values. Duplicate pairs were verified through concordance of genotypes for 65 SNPs on the Methylation 450 chip.

*PCA for MWAS*

Because our sample included salivary DNA samples, we were unable to adjust for cell-type composition using a blood-sample-based reference. Instead, we conducted principal components analysis (PCA) on genomewide methylation as follows: We selected 287,720 CpG sites with SD ≥ 0.02 across the entire sample and normalized the beta values for each site to mean = 0, SD = 1, creating an m × n matrix X, where m is the number of markers and n the number of samples. The eigenvectors from the matrix X′X/(m – 1), an n × n matrix, were obtained using the eigen() function in R, to be used as PC covariates in methylome-wide association studies (MWAS). We regressed our SSD outcomes on each of the first 20 PCs, and included significantly associated PCs in MWAS. Phenotypes were adjusted for between one and four PCs.

*Methylation-QTL (meQTL) Analysis*

We conducted a targeted cis-methylation QTL analysis over 521 CpG sites within 50 kilobasepairs (kb) of 162 candidate SNPs (Supplemental Table 2), using Matrix eQTL ^22^ to find the effect of genotype on extent of methylation in a sample of 597 individuals with both epigenetic and imputed genotype data. All pairs of SNPs and CpG sites within 100 kb were considered to be in cis. Methylation was expressed as M values, where M = log(β/(1 – β)), which extends the range of possible values to (–∞,∞), making the values suitable as an outcome for linear regression.

**Endophenotype-based polygenic risk score methods**

We generated polygenic risk scores in the European subset of the CFSRS where genotype data, as well as clinical group data (no disorder, SSD only, language impairment (LI) only, SSD+LI, CAS) were available. Risk scores relied upon association statistics (beta coefficients) from our CFSRS GWAS and were constructed using PLINK 1.9^23^ (clump and score functions). Regions were considered if at least one variant in the region met the threshold for inclusion as a risk variant (P < 0.001). Clumping of variants was done in selected regions around the variant showing the strongest association in the region, removing other variants in linkage disequilibrium (r^2^> 0.5). We used a linear mixed model to model the relationship between polygenic risk score and clinical group, controlling for sex and familial relationship (based on family ID*)*. Nested model comparison (the full model with clinical group included versus the reduced model with clinical group removed) using the chi-squared test was implemented to determine if clinical group explained a significant amount of variability in polygenic risk.

**Transcriptome-wide association study**

We applied the EpiXcan pipeline ^24^ to train gene expression predictors in human brain tissue. For genotypes and gene expression, we used psychENCODE data from the dorsolateral prefrontal cortex (DLPFC)^25^. We restricted our analysis to 924 Caucasian samples. We initially computed eQTL summary statistics using the R package MatrixEQTL^22^, followed by estimation of SNP priors through the qtlBHM Bayesian hierarchical model ^26^ using the Roadmap Epigenomics Project chromatin states for DLPFC (‘BRN_DL_PRFRNTL_CRTX’). In total, 363,955 predictors for 18,425 genes were recruited in the EpiXcan psychENCODE model. We then applied the S-PrediXcan method ^27^ using the EpiXcan psychENCODE model as well as the SNP covariance matrix on the GWAS summary statistics. These analyses were based on genome-wide association results from two phenotypes from our GWAS, TWS and Elision; these traits were chosen because they had the greatest number of unique significantly-associated loci. Detailed results are in Supplemental Tables 5 and 6.

**GWAS methods for ALSPAC**

Genotype QC was performed previously by ALSPAC^28^. We restricted our ALSPAC sample to unrelated individuals by randomly removing one from a pair of twins, when applicable. PCs were generated using Hail 0.1 software, to accommodate the format of files obtained from ALSPAC, using a standard PCA approach^29^. In generating the PCs we first removed long range LD regions and restricted to variants with a MAF > 0.01, an imputation quality score of > 0.95 and variants not in LD (r^2^ < 0.2; following the same process as with PLINK’s --indep-pairwise default procedure). Genetic association testing was performed using linear regression in Hail 0.1 when outcome measures were continuous and using logistic regression in Hail 0.1 when outcome measures were binary. We restricted our analyses to variants with a MAF > 0.01 and an imputation quality score of > 0.6; we used a lower MAF threshold because we hypothesized that causal variants might be rarer in a population-based cohort compared to a cohort that was ascertained through a trait of interest. Covariates adjusted for included sex and the first two PCs. Age was not a consideration as ALSPAC is a longitudinal birth cohort study and age differences were negligible for any given measure.

1	Robbins, J. & Klee, T. Clinical assessment of oropharyngeal motor development in young children. *Journal of Speech and Hearing Research* **52**, 271-277 (1987).

2	Fletcher, D. (C.C. Publications, Inc., Tigard, OR, 1977).

3	Gardner, M. (Academic Therapy Publications, Novato, CA, 1990).

4	Dunn, L. & Dunn, L. (American Guidance Service, Inc, Circle Pines, MN, 1997).

5	Catts, H. Speech production/phonological deficits in reading disordered children. *Journal of Learning Disabilities* **19**, 504-508 (1986).

6	Lewis, B. A. *et al.* Speech and language skills of parents of children with speech sound disorders. *Am J Speech Lang Pathol* **16**, 108-118 (2007).

7	Wagner, R. T., J; Rashotte, C; Pearson, NA. (Pearson, London, England, 2013).

8	Denckla, M. R., RG;. Rapid automatized naming of pictured objects, colors, letters and numbers by normal children. *Cortex* **10**, 186-202 (1974).

9	E, S., Wiig, E. & Secord, W. *Clinical evaluation of language fundamentals-Revised*. (The Psychological Corporation, 1987).

10	Newcomer, P. & Hammill, D. *Test of language development - Primary, Second Edition*. (Pro-Ed., 1988).

11	Woodcock, R. (American Guidance Service, Circle Pines, MN, 1987).

12	Wechsler, D. (The Psychological Coporation, San Antonio, TX, 1991).

13	Lewis, B. A. *et al.* Literacy outcomes of children with early childhood speech sound disorders: impact of endophenotypes. *J Speech Lang Hear. Res* **54**, 1628-1643 (2011).

14	Wellman, R. L. *et al.* Narrative ability of children with speech sound disorders and the prediction of later literacy skills. *Lang Speech Hear. Serv. Sch* **42**, 561-579 (2011).

15	*Nonsense word repetition task*.

16	Nunes, T. B., P; Olsson, J; . Learning Morphological and Phonological Spelling Rules: An Interventinoal Study. *Scientific Studies of Reading* **7**, 289-307 (2009).

17	Novembre, J. *et al.* Genes mirror geography within Europe. *Nature* **456**, 98-101, doi:10.1038/nature07331 (2008).

18	Das, S. *et al.* Next-generation genotype imputation service and methods. *Nat Genet* **48**, 1284-1287, doi:10.1038/ng.3656 (2016).

19	Howie, B. N., Donnelly, P. & Marchini, J. A flexible and accurate genotype imputation method for the next generation of genome-wide association studies. *PLoS. Genet* **5**, e1000529 (2009).

20	Assenov, Y. *et al.* Comprehensive analysis of DNA methylation data with RnBeads. *Nat. Methods* **11**, 1138-1140 (2014).

21	Teschendorff, A. E. *et al.* A beta-mixture quantile normalization method for correcting probe design bias in Illumina Infinium 450 k DNA methylation data. *Bioinformatics* **29**, 189-196, doi:10.1093/bioinformatics/bts680 (2013).

22	Shabalin, A. A. Matrix eQTL: ultra fast eQTL analysis via large matrix operations. *Bioinformatics* **28**, 1353-1358, doi:10.1093/bioinformatics/bts163 (2012).

23	Purcell, S. *et al.* PLINK: a tool set for whole-genome association and population-based linkage analyses. *Am J Hum Genet* **81**, 559-575 (2007).

24	Zhang, W. *et al.* Integrative transcriptome imputation reveals tissue-specific and shared biological mechanisms mediating susceptibility to complex traits. *Nature communications* **10**, 3834, doi:10.1038/s41467-019-11874-7 (2019).

25	Wang, D. *et al.* Comprehensive functional genomic resource and integrative model for the human brain. *Science* **362**, doi:10.1126/science.aat8464 (2018).

26	Li, Y. I. *et al.* RNA splicing is a primary link between genetic variation and disease. *Science* **352**, 600-604, doi:10.1126/science.aad9417 (2016).

27	Barbeira, A. N. *et al.* Exploring the phenotypic consequences of tissue specific gene expression variation inferred from GWAS summary statistics. *Nature communications* **9**, 1825, doi:10.1038/s41467-018-03621-1 (2018).

28	Eicher, J. D. *et al.* Genome-wide association study of shared components of reading disability and language impairment. *Genes Brain Behav* **12**, 792-801, doi:10.1111/gbb.12085 (2013).

29	Price, A. L. *et al.* Principal components analysis corrects for stratification in genome-wide association studies. *Nat. Genet* **38**, 904-909 (2006).

**SUPPLEMENTAL TABLES**

**Supplemental Table 1. Descriptive statistics for CFSRS measures**

| Test | N | Mean age (SD) [range] | % Female | % Lang | % SSD | % CAS |
| --- | --- | --- | --- | --- | --- | --- |
| NSW_PPC | 431 | 14.0 (12.4) [4, 64] | 45% | 30% | 54% | 11% |
| NSW | 431 | 14.0 (12.4) [4, 64] | 45% | 30% | 54% | 11% |
| MSW_PPC | 432 | 14.0 (12.4) [4, 64] | 45% | 30% | 55% | 11% |
| MSW | 432 | 14.0 (12.4) [4, 64] | 45% | 30% | 55% | 11% |
| PPVT | 399 | 12.5 (12.0) [2.5, 64] | 44% | 32% | 57% | 12% |
| EOWPVT | 364 | 10.4 (9.6) [2.5, 64] | 42% | 33% | 59% | 13% |
| WRID | 399 | 16.4 (11.7) [5, 64] | 45% | 29% | 53% | 10% |
| WRAT | 398 | 16.4 (11.7) [5, 64] | 45% | 29% | 53% | 10% |
| TWS | 298 | 10.3 (2.4) [5, 18] | 40% | 33% | 59% | 12% |
| Elision | 299 | 10.4 (3.7) [4, 22] | 40% | 35% | 62% | 14% |
| RAN | 309 | 9.0 (3.9) [4, 23] | 40% | 33% | 62% | 14% |
| CELF_Receptive | 325 | 7.4 (3.1) [3, 18] | 40% | 34% | 62% | 13% |
| CELF_Expressive | 325 | 7.4 (3.1) [3, 18] | 40% | 34% | 62% | 13% |
| Fletcher | 419 | 13.5 (12.7) [2.5, 64] | 45% | 29% | 55% | 11% |
| WIAT_LC | 165 | 16.2 (11.9) [7, 51] | 44% | 27% | 42% | 3.70% |
| WIAT_RC | 166 | 16.0 (11.9) [7, 51] | 45% | 28% | 42% | 3.60% |

**Supplemental Table 2 – Results of methylation analysis of candidate gene regions**

This file is supplied as a separate excel file

**Supplemental Table 3. Descriptive statistics for ALSPAC sample**

| Sample size (union across all tests) | 9658 |
| --- | --- |
| Age range | [5, 9] |
| Female N (%) | 4773 (49%) |
| Speech problems (parent reported) | 6% |

**Supplemental Table 4. Correspondence between CFSRS and ALSPAC measures**

| **CFSRS measure** | **Corresponding ALSPAC measure(s)** | **Age given** | **Sample size for ALSPAC** |
| --- | --- | --- | --- |
| Multisyllabic word repetition (MSW) | Repetition of “buttercup” and “dinosaur” (**MWR)** | 5 | 711 |
| Nonsense word repetition (NSW) | Children’s test of non-word repetition (**CNrep5**)  ----  12 items from Children’s test of non-word repetition (**CNrep8**) | 5  8 | 680  5860 |
| Woodcock real-word reading (WRID) | WORD single word reading (**WORD**)  ----  Neale analysis of reading ability (**NARA**) – reading comprehension (**NARA-C**) and reading accuracy subtests **(NARA-A**) | 7 |  |
| Woodcock nonsense word reading (WRAT) | Single word nonword reading **(ALSPACread)** | 9 | 6136 |
| Test of written spelling (TWS) | Single word spelling (**ALSPACspell7**)  -----  Single word spelling (**ALSPACspell9**) | 7  9 | 6203  6137 |
| CELF receptive (CELF_R) | WOLD comprehension (**WOLD-C**) | 8 | 5865 |
| CELF expressive (CELF_E) | WOLD expression (**WOLD-E**) | 8 |  |
| Expressive one word vocabulary (EOWPVT) | WOLD expression – one word vocabulary test | 8 | 5840 |
| Peabody picture vocabulary (PPVT) | WISC vocabulary (**WISC-V**) | 8 | 5844 |

**Supplemental Table 5. PsychEncode EpiXcan method using Meta-analysis results of Elision GWAS**

| **zscore** | **effect_size** | **pvalue** | **var_g** | **pred_perf_r2** | **pred_perf_pval** | **pred_perf_qval** | **n_snps_used** | **n_snps_in_cov** | **n_snps_in_model** | **fdr** |
| --- | --- | --- | --- | --- | --- | --- | --- | --- | --- | --- |
| -4.39 | -0.70 | 1.11E-05 | 0.015 | 0.014 | 3.67E-04 | 8.12E-04 | 14 | 17 | 15 | 0.14 |
| 4.01 | 0.13 | 5.97E-05 | 0.342 | 0.411 | 6.52E-117 | 3.53E-115 | 23 | 29 | 29 | 0.75 |
| -4.01 | -0.78 | 6.17E-05 | 0.009 | 0.021 | 1.13E-05 | 2.81E-05 | 7 | 17 | 16 | 0.77 |
| 3.97 | 0.97 | 7.27E-05 | 0.006 | 0.006 | 2.26E-02 | 4.35E-02 | 11 | 22 | 17 | 0.91 |
| -3.65 | -0.68 | 2.59E-04 | 0.011 | 0.024 | 1.80E-06 | 4.71E-06 | 11 | 18 | 12 | 1.00 |
| -3.63 | -0.33 | 2.89E-04 | 0.043 | 0.065 | 2.63E-15 | 1.13E-14 | 30 | 63 | 63 | 1.00 |
| 3.57 | 0.65 | 3.51E-04 | 0.008 | 0.010 | 2.89E-03 | 5.98E-03 | 14 | 23 | 22 | 1.00 |
| -3.51 | -0.49 | 4.52E-04 | 0.019 | 0.051 | 4.23E-12 | 1.55E-11 | 18 | 22 | 19 | 1.00 |
| 3.50 | 1.14 | 4.73E-04 | 0.004 | 0.017 | 7.02E-05 | 1.64E-04 | 2 | 2 | 2 | 1.00 |

**Supplemental Table 6. PsychEncode EpiXcan method using Meta-analysis results of TWS GWAS**

| **zscore** | **effect_size** | **pvalue** | **var_g** | **pred_perf_r2** | **pred_perf_pval** | **pred_perf_qval** | **n_snps_used** | **n_snps_in_cov** | **n_snps_in_model** | **fdr** |
| --- | --- | --- | --- | --- | --- | --- | --- | --- | --- | --- |
| -4.18 | -1.34 | 2.88E-05 | 0.009 | 0.010 | 2.07E-03 | 4.32E-03 | 8 | 30 | 24 | 0.36 |
| 3.96 | 0.45 | 7.64E-05 | 0.077 | 0.104 | 4.50E-24 | 2.77E-23 | 23 | 29 | 29 | 0.95 |
| -3.57 | -0.36 | 3.55E-04 | 0.111 | 0.166 | 4.68E-39 | 4.67E-38 | 12 | 13 | 13 | 1.00 |
| -3.52 | -2.17 | 4.28E-04 | 0.003 | 0.014 | 3.28E-04 | 7.29E-04 | 3 | 6 | 3 | 1.00 |
| -3.51 | -0.21 | 4.50E-04 | 0.216 | 0.308 | 3.78E-80 | 9.68E-79 | 4 | 6 | 4 | 1.00 |
| 3.49 | 1.69 | 4.85E-04 | 0.004 | 0.015 | 2.23E-04 | 5.02E-04 | 4 | 4 | 4 | 1.00 |
| -3.41 | -0.24 | 6.49E-04 | 0.191 | 0.229 | 4.18E-56 | 6.37E-55 | 55 | 100 | 89 | 1.00 |
| 3.33 | 0.53 | 8.69E-04 | 0.041 | 0.161 | 4.38E-38 | 4.25E-37 | 21 | 39 | 34 | 1.00 |
| 3.33 | 1.04 | 8.77E-04 | 0.009 | 0.015 | 1.92E-04 | 4.35E-04 | 15 | 25 | 25 | 1.00 |
| -3.32 | -0.33 | 8.99E-04 | 0.079 | 0.107 | 2.87E-25 | 1.85E-24 | 20 | 33 | 29 | 1.00 |

**Supplemental Table 7 – Association results from regions identified from published GWAS of reading and language phenotypes**

This file is provided as a separate excel file named Supplemental Table 7

**Supplemental Figures**

**Supplemental Figure 1. Locus zoom plots and functional annotation for most signfiicant findings.** Supplemental Figure 1 shows association results for the top loci. P-values displayed are for CFSRS and are for the test for which the top SNP was observed. Circles show P-values for SNP associations and triangles show P-values for methylation associations (specifically those for which the top SNP is a meQTL for). The larger plot shows the top SNP for each region +/- 200 kb. The window highlights the region that spans significant association results (P≤ 1×10-5 in CFSRS) in any CFSRS test. A. IFI16 region (window spans chr1:159001292-159028378); B. NFKBIA region (window spans chr14:35770806-35846092); C. DACT1 region (window spans chr14:59210335-59221002); D. SETD3 region (window spans chr14:99858970-99942692). E. MON1B region (window spans chr16:77231207-77248555).

**Suuplemental Figure 1a**

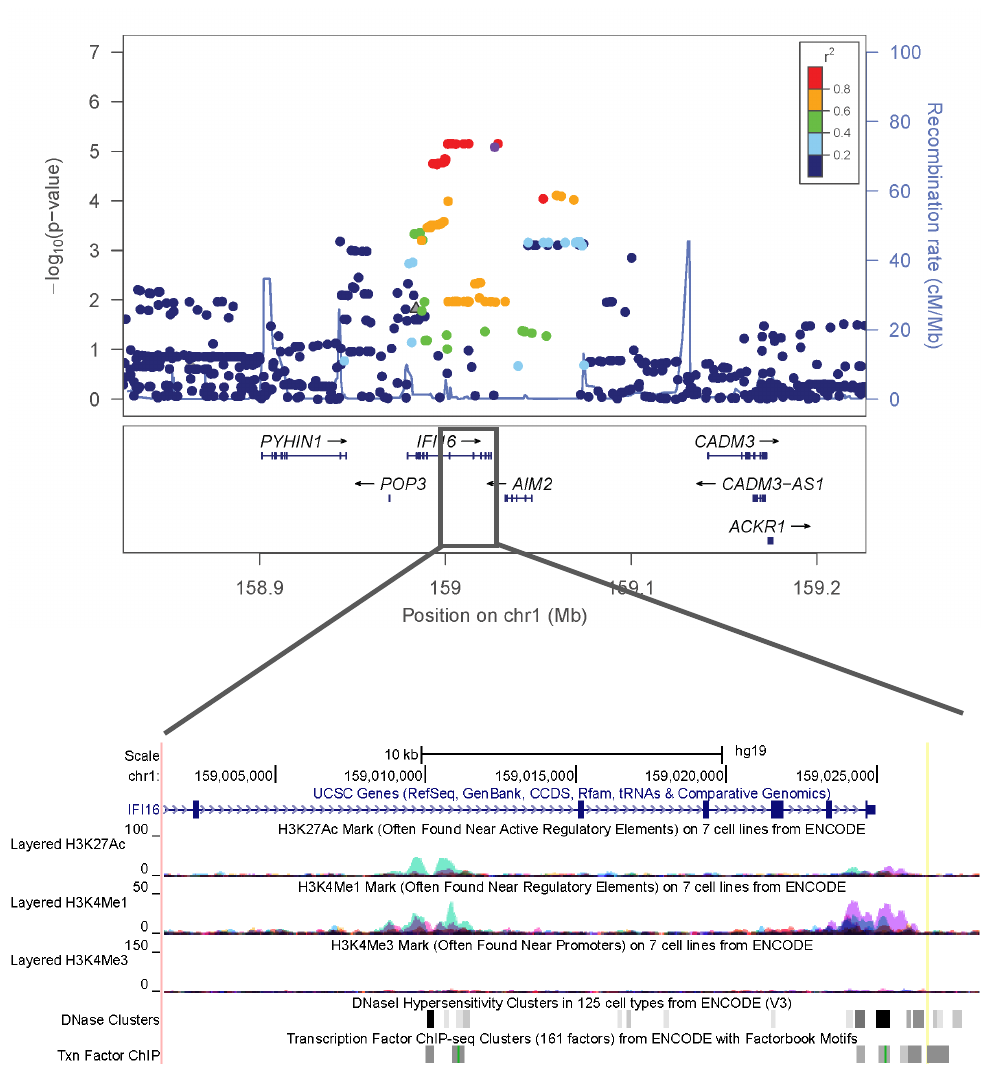

**Supplemental Figure 1b.**
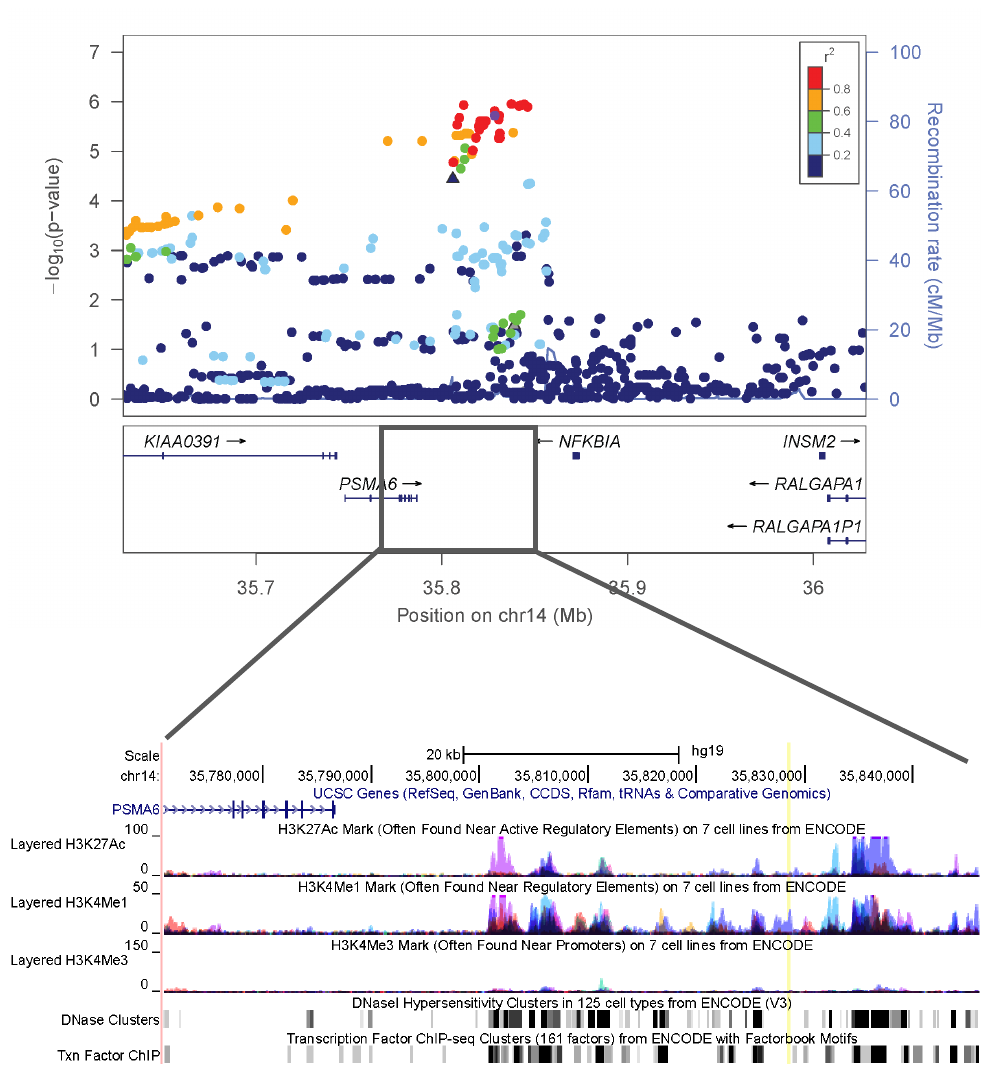

**Suuplemental Fig 1c**
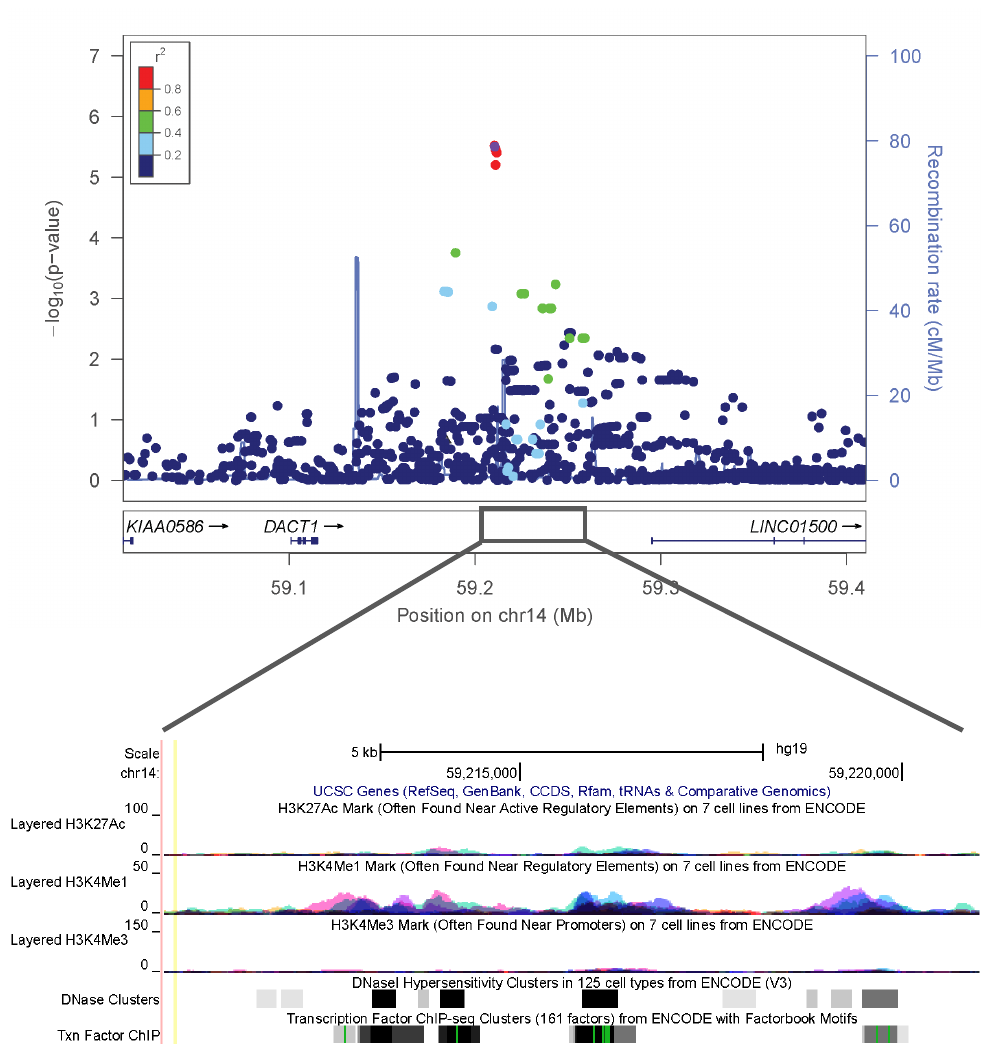
**Suuplemental Fig 1d**
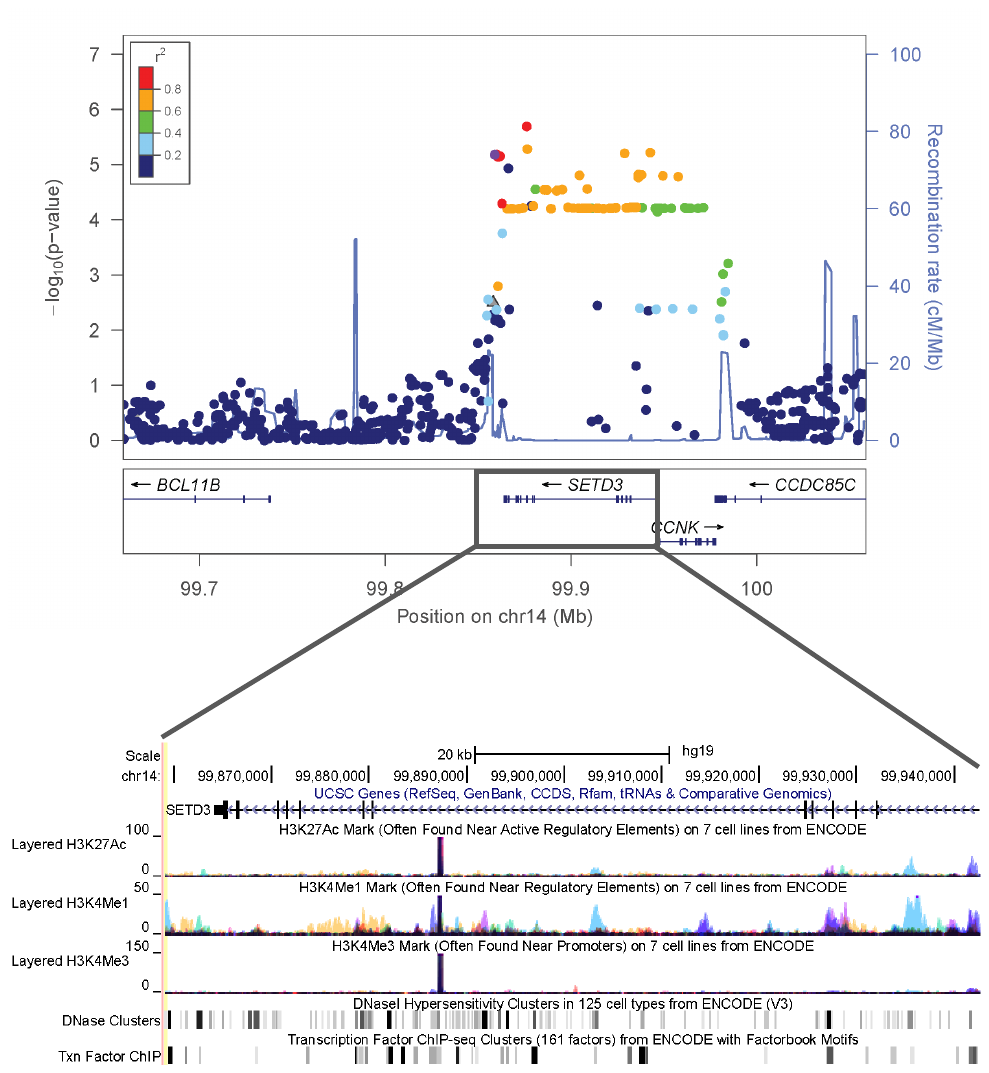

**Supplemental Fig 1e.**

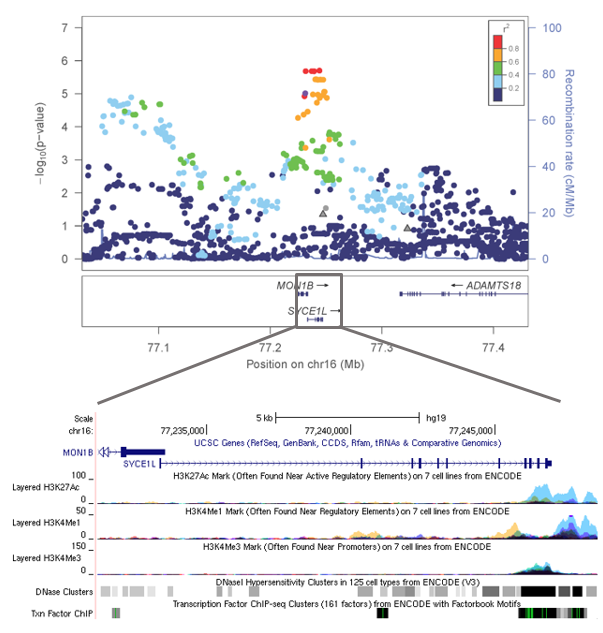

**Supplemental Figure 2. Clustering of Significant Variants (P < 0.01) among Known Speech Genes across CFSRS Tests**

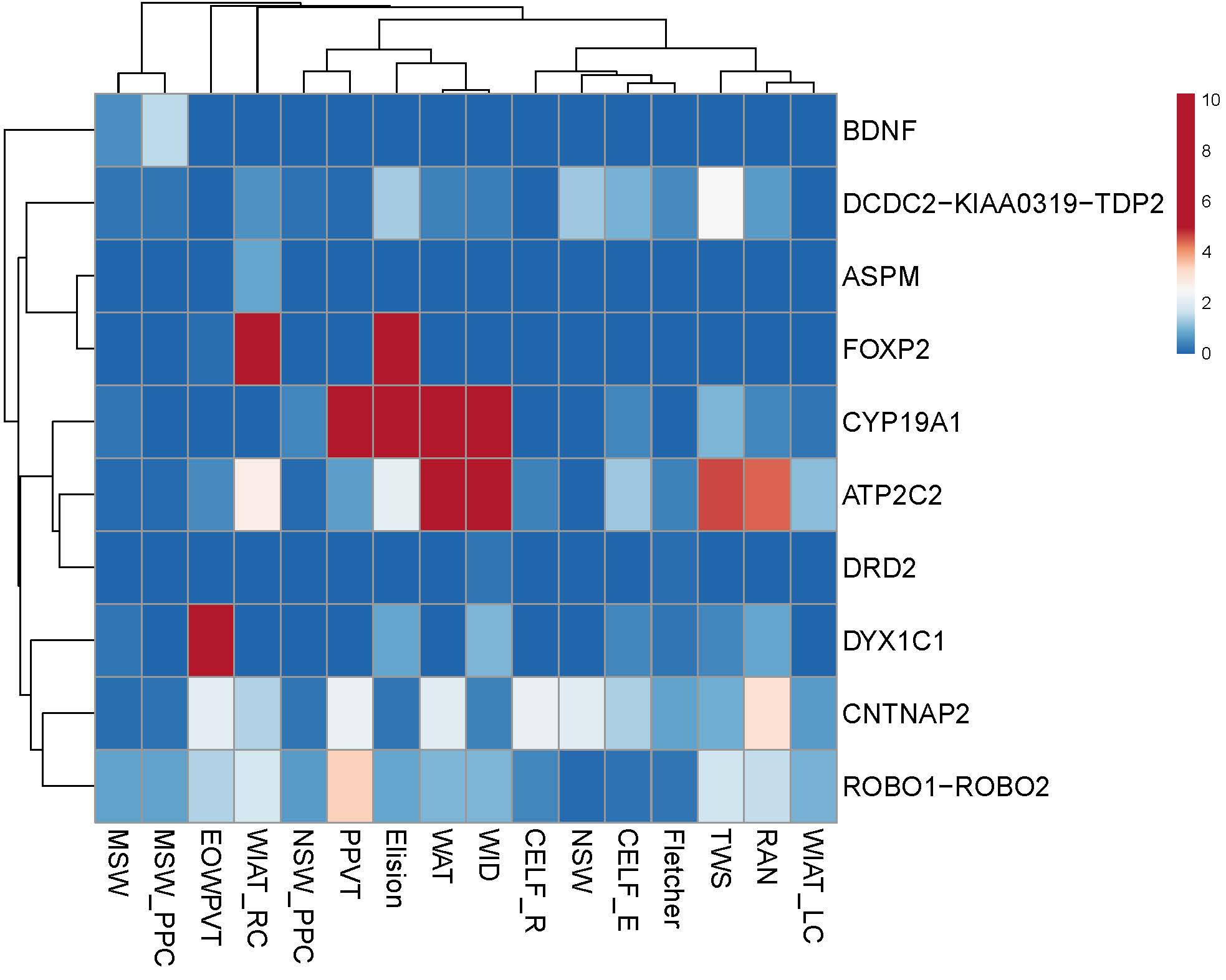

**Supplemental Figure 3 – in separate pdf**

LocusZoom plots of candidate genes where at least one trait had a SNP significant at p < 10^-4^

**Supplemental Figure 4. Clustering of Significant Variants (P < 0.01) among Known Speech Genes across ALSPAC Tests**

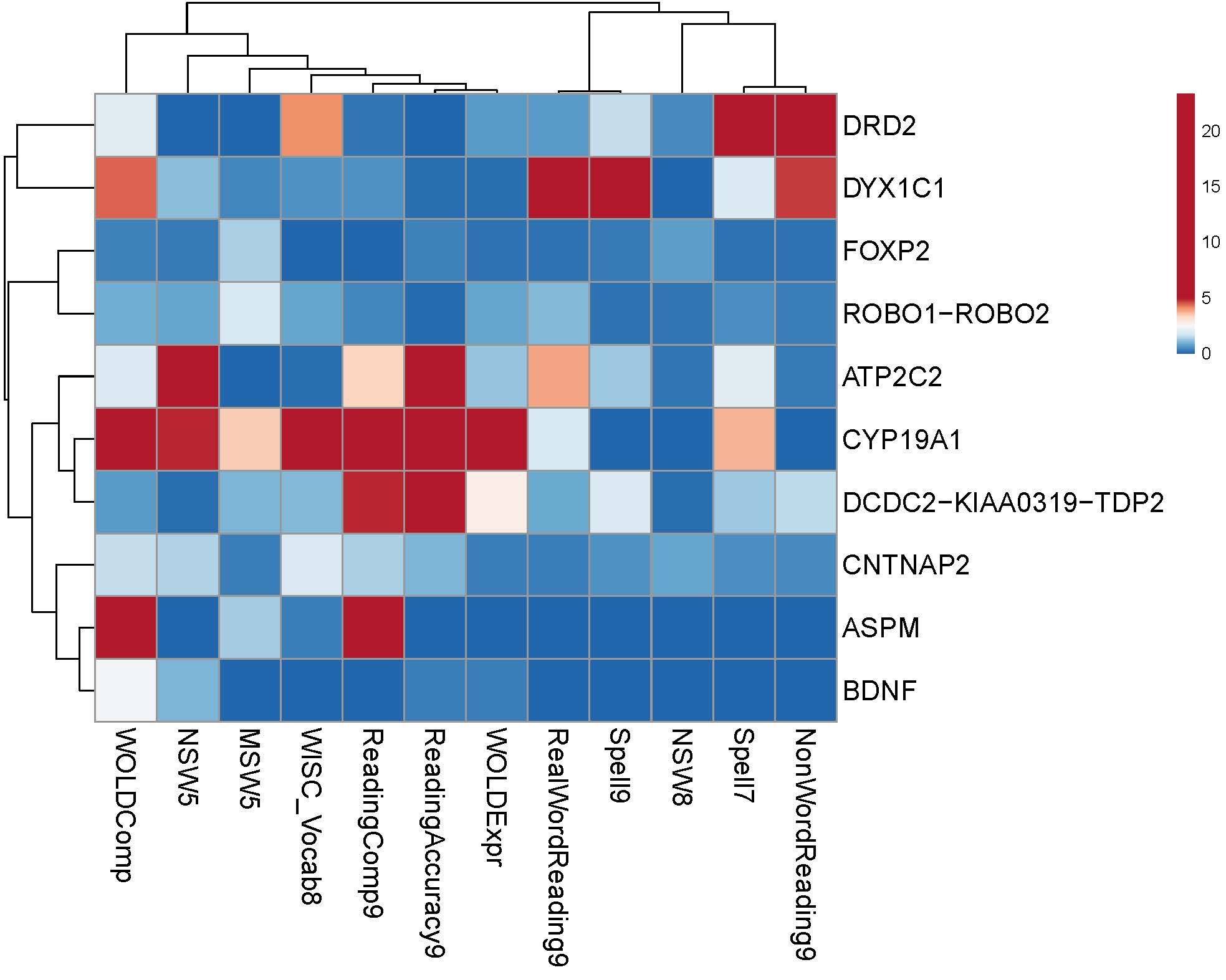

**Supplemental Figure 5. Polygenic Risk score across all individual measures**

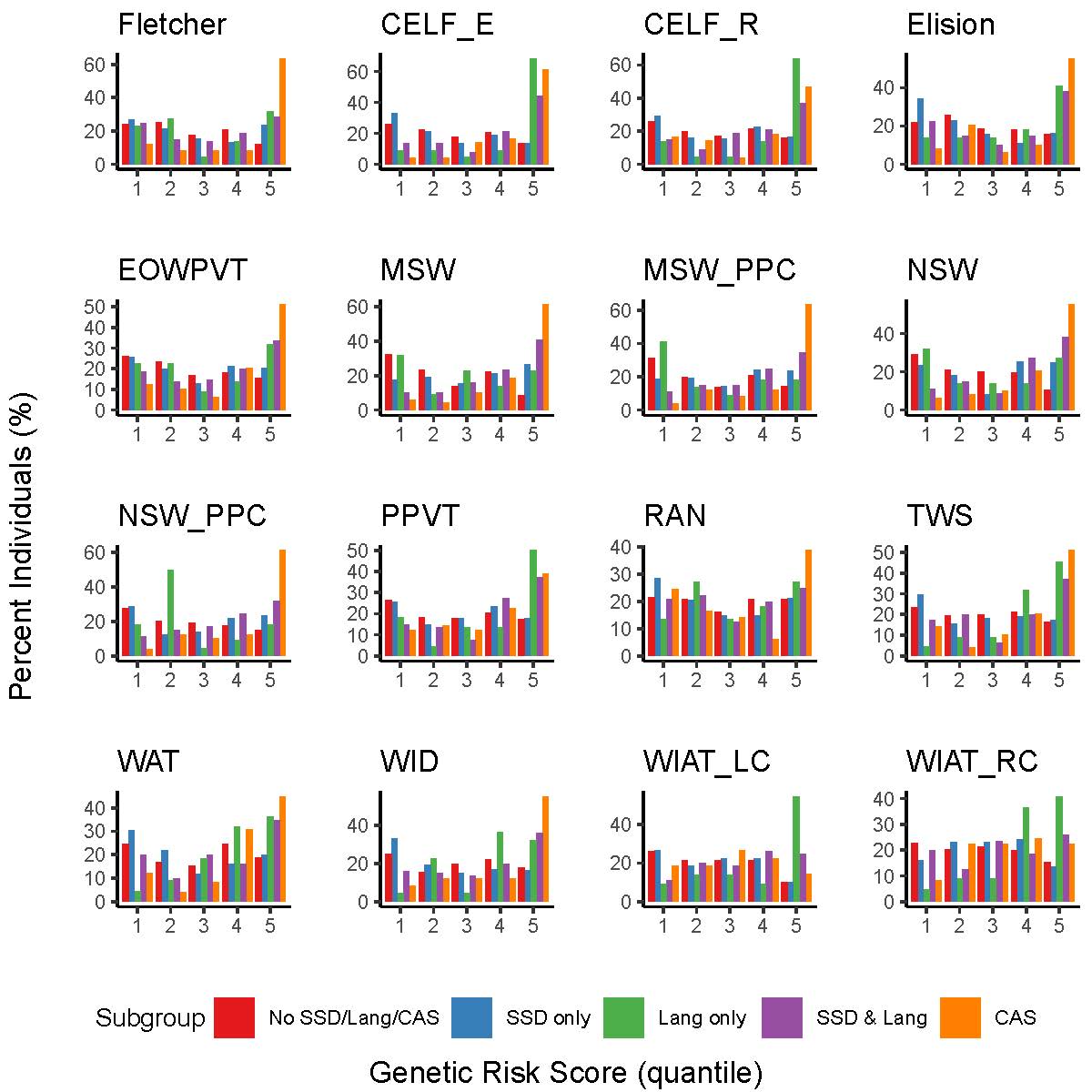
